# Clinical medicine admission cutoffs in China, 2017-2026: a coverage-sensitive national longitudinal analysis

**DOI:** 10.64898/2026.11.25.26368769

**Authors:** Hang Zhou, Xiuyuan Li

## Abstract

**Context.:** Clinical medicine cutoffs locate published score or rank boundaries at filing or admission. China’s 31 provincial systems support a ten-year national analysis of these boundaries and their career context.

**Methods.:** Undergraduate clinical medicine cutoffs were identified in examination cohorts across the 31 provincial systems from 2017 to 2026. The estimand was the unweighted mean of provincial median school cutoffs. Filing and admission stages were not consistently distinguished. Schools and pathways were compared at matched endpoints under alternative school identities. Comparisons beginning at the 2023 maximum were post hoc and descriptive.

**Results.:** Of 310 province-year cells, 262 met the three-school threshold. Among 407 province– school pairs observed throughout the decade, mean percentiles were 76.23 in 2017, 86.06 in 2023 and 76.51 in 2026. Across 754 endpoint pairs, the matched-school change was +3.02 points. School and province coverage expansion changed the estimate by −6.44 and −0.49 points, yielding −3.90. In 2023–2025, provincial medians moved later in 25 of 29 provinces and earlier in four; mean change was −2.31 points (2,084 pairs; 95% interval −3.36 to −1.25). In post hoc 2023–2026, the corresponding values were 25 and four of 29 provinces, −5.65 points, and 1,345 pairs (95% interval −7.76 to −3.50). Four school-identity definitions yielded −5.65 to −5.43.

**Conclusions.:** The observed maximum in the 15-province fixed panel was in 2023; cutoffs moved later by 2026. This recent direction was stable under alternative school identities and matched examination populations, while its magnitude remained coverage-sensitive. Long hours, a relative-earnings ratio that declined after 2017, lengthy training and varied recruitment requirements describe career context but do not identify causes. Cutoffs describe published filing or admission boundaries, not applicant demand or future physician supply.

**Practice Points:**

Longitudinal cutoffs can reverse direction when school coverage changes.
Matching schools within provinces separates composition from within-school movement.
The recent movement was stable across school-identity and examination-category checks.
Career conditions describe context but do not identify causes of cutoff movement. Admission boundaries should not be read as applicant demand or future physician supply.

## Introduction

When a medical programme admits a candidate further down the examination ranking, the cutoff locates an observed competition boundary. Applicant preferences, available places, eligibility rules and allocation procedures all shape that position. A national series can therefore show how the boundary changed and locate the training and career conditions around that change. The design of the selection system also shapes how candidates enter medicine. ^1,2^

Clinical medicine in China includes five-year, integrated five-plus-three, six-year and eight-year routes. These routes differ in subject requirements and in their transition to postgraduate training. Recruitment schedules from leading Chinese tertiary hospitals commonly distinguish between master’s and doctoral qualifications. In a 2022 survey of 113 medical schools, clinical students with job-search experience most often cited educational qualifications as a difficulty. ^3^ These graduate experiences describe the endpoint of a training pathway that applicants may begin to evaluate much earlier.

Public discussion of medicine in China has increasingly centred on long hours, extended training and uncertain returns. National surveys of practising physicians provide a second view of that pathway. Physicians in tertiary public hospitals have reported long patient-service weeks and a substantial intention to leave the profession. Entrant surveys also show that most students expected further study before employment. A national student survey recorded workload and training duration as prominent reasons for rejecting medicine. These conditions form the public backdrop against which families compare medicine with other courses. Measurements made at later career stages are used in the Discussion to interpret the admission trends.

Earlier analyses documented an upward phase before the present observation window. Across 21 clinical medicine programmes, admission scores rose from a relatively moderate baseline in 2013–2015. ^4^ A comparison of 12 comprehensive universities reported that the average admission rank moved earlier between 2011 and 2016. ^5^ Other work examined average entry scores in 14 Chinese five-year programmes with sufficient data for a 2013–2016 comparison. ^6^ A nationwide study also counted 70,480 planned undergraduate clinical medicine places at 180 institutions in 2018. ^7^ The present study covers 2017–2026, measures each cutoff within its provincial candidate distribution and follows continuing schools into the recent reversal. It asks how candidate-referenced clinical medicine cutoffs changed across the decade and within continuing schools and training pathways. A methodological subquestion concerns their sensitivity to school and province coverage. Career conditions and comparable admission systems are then examined as context in the Discussion. They do not form additional outcome series.

## Methods

### Records and outcomes

This retrospective longitudinal study used published admission records from 31 mainland Chinese provincial examination systems for the 2017–2026 cycles. Province refers to the system through which candidates applied. Institution and vocational programme registers were linked by programme title, admission batch and route description to identify ordinary undergraduate clinical medicine. Vocational programmes, unresolved degree levels, cooperative programmes and explicitly identified special or service-linked routes were excluded. The sources reported a minimum score or rank at filing or admission. Filing is the stage at which a candidate’s record is forwarded to a school. Because sources did not consistently identify that stage across provinces and years, the primary outcome is reported as a filing-or-admission cutoff.

The primary outcome located each cutoff within the reported candidate distribution for its province, year and examination category. Its percentile was 100 × [1 − (r − 1)/N], where r is the linked rank (1 is highest) and N is the number of candidates in that distribution. The linked rank was obtained from the cumulative count at the cutoff score or from a reported rank. When a cutoff score represented a tied group, the cumulative count at that score, corresponding to the tie-end rank, was used. Scores beyond the published table were assigned the corresponding endpoint rank. Higher percentiles indicate earlier positions. A secondary measure ranked the cutoff among recorded programme cutoffs in the same province, year, examination category and admission batch.

### Longitudinal comparisons

For each year, eligible records were first medianed within each school and examination province. School medians were then medianed within each province. The national estimand was the unweighted mean of these provincial medians among observed eligible schools. Each province therefore received equal weight, and schools were equally weighted within a province after the school-level median. The annual estimate included only provinces contributing at least three schools. The calculation was repeated using schools observed throughout all ten years, with at least three continuing schools per province. Regional variation was examined in this fixed panel, and the national comparison was repeated with each province omitted in turn. Schools present at both the 2023 and 2026 endpoints were also compared. A stricter specification required presence in every intervening year. As an aggregation check, each school median was weighted by its number of contributing programme records before the provincial mean was taken. All 310 province-year cells were reported, including years with insufficient coverage.

Three windows entered the coverage decomposition: 2017–2026, 2023–2026 and 2025–2026. Within each window, schools were matched within province at the two endpoints. Windows beginning in 2023 start at the observed maximum in the annual point estimates. Because that endpoint was selected after inspecting the annual series, comparisons beginning in 2023 are post hoc and descriptive. An additional endpoint-matched comparison covered 2023–2025. It used the same province-equal aggregation, three-school threshold and province-resampling procedure to assess whether the recent direction predated the dominant 2026 source transition. For each of the three coverage-decomposition windows, the comparison was successively restricted to common provinces, provinces with at least three common schools and the common schools themselves. Differences between these estimates give an order-dependent decomposition. Minimum school counts of 1, 3, 5 and 10 were tested in sensitivity analyses, as were three approaches to duplicate records (supplement).

Province-resampling intervals describe the stability of observed provincial estimates under province resampling. All schools within a resampled province were retained. The reported 95% interval used the percentile method with 50,000 replicates and random seed 20261123 (Table S1). It is not a sampling interval from all possible provincial systems to a superpopulation. It also does not account for post hoc endpoint selection, nonrandom coverage or measurement error in the source records.

The primary school identity used the institution name recorded in each source. The 2023–2026 comparison was repeated using the national school code alone. It was also repeated using code followed by a normalised name when code was absent, and using code followed by the source match name. Records without a code were excluded only from the code-only specification. The comparison was also repeated after stratifying each school by examination category. Science-track labels, humanities-track labels, Mongolian-language science, comprehensive systems and unlabelled records were kept separate. Category stratification yielded 1,344 matched pairs in the same 29 provinces. One unlabelled Heilongjiang 2024 unit contained two reference denominators and was excluded from this comparison. A further 105 school-year units combined more than one original category label (Supplementary Tables S2–S6).

Adjacent-year comparisons required both percentile measures at both endpoints and at least three matched schools per province. Change was compared at each school and in the provincial median across those schools. Directional agreement counted pairs that moved earlier on both measures or later on both. Ties remained in the denominator. For 2025–2026, the dominant exact source tier and broader source family were recorded for each province–school–year. "Mixed" was assigned when tiers tied. Transitions were tabulated alongside the matched cutoff change (Supplementary Tables S7–S9).

### Training pathways and examination populations

For each province-year, the contributing files, reference distributions, school count, quantity-field definitions and usable-cutoff availability were inventoried. Reported counts with unclear definitions were retained in a separate field. Independently dated plans, filing-stage allocations, forwarded candidates, completed admissions and first-programme choices were coded separately. The supplement documents source-level checks against original records.

A supplementary source audit documented school-level plan evidence for 100 province-year cells. The audit retained dated sources, original field labels and record hashes, and reconciled conflicting quantities without constructing a provincial capacity total.

Five-year, integrated five-plus-three, six-year and eight-year routes were compared where the recorded title or duration allowed classification. At each endpoint, schools with at least one common pathway were retained. For a given set of schools and provinces, all classifiable records were used first. The comparison was then limited to pathways present at both endpoints, with each shared pathway weighted equally within its school through the median of pathway medians. The analysis was repeated using explicit durations alone. Programmes explicitly labelled five-year and five-plus-three were compared within the same school, province, year, examination category, admission batch and candidate denominator.

To distinguish movement in the underlying rank from change in the reference population, schools were matched within examination categories across endpoints. Synonymous category labels were grouped, while traditional science and new physics routes remained distinct. Contributions from rank and reference-population change were calculated by averaging the two possible replacement orders for the starting rank and denominator. The supplement gives the aggregation procedure, sensitivity to original category labels and provincial coverage.

### Historical career context and recruitment

Contextual evidence covered dated national surveys, payroll and registry studies, hospital recruitment schedules and student surveys. ^8–23^ Sources were inventoried by period, geographic coverage, sampling frame and denominator. No continuous 2020– 2026 national series of weekly hours or earnings was available. Records from different years and frames were therefore retained as separate snapshots and used only in the Discussion or supplement. The recruitment search covered the 100 hospitals in the 2023 Fudan ranking for notices dated 1 July to 20 November 2026. It also covered one medical school’s complete employment board for the 2024–2026 graduating classes and a separate 2025 recruitment schedule from one national hospital group. ^8–12^ Recruitment records were retained only when employer, department and recruitment channel could be identified. Degree requirements, university credentials, publication criteria and disclosed pay were then coded using the supplement rules. Only readable schedules, non-senior clinical posts and explicitly stated degree requirements entered the supporting career-condition figure. Schedule lines and advertised places were counted separately. Leading-hospital and national-group frames were never pooled. These records provide context and do not form a second outcome series.

## Results

### National trajectory and coverage

Of 36,866 records screened across 31 provincial examination systems, 34,484 met the eligibility criteria for ordinary undergraduate clinical medicine. Thirty provinces contributed at least once to the annual estimates (Figure 1b). The three-school threshold was met in 262 of 310 possible province-year cells, accounting for 34,345 records. Twenty provinces contributed in 2017 and 30 in 2026. Recent annual comparisons included 28–30 of the 31 systems in every year from 2021 to 2026. Fifteen provinces and 407 province–school pairs were followed throughout the ten years.

**Figure 1.**
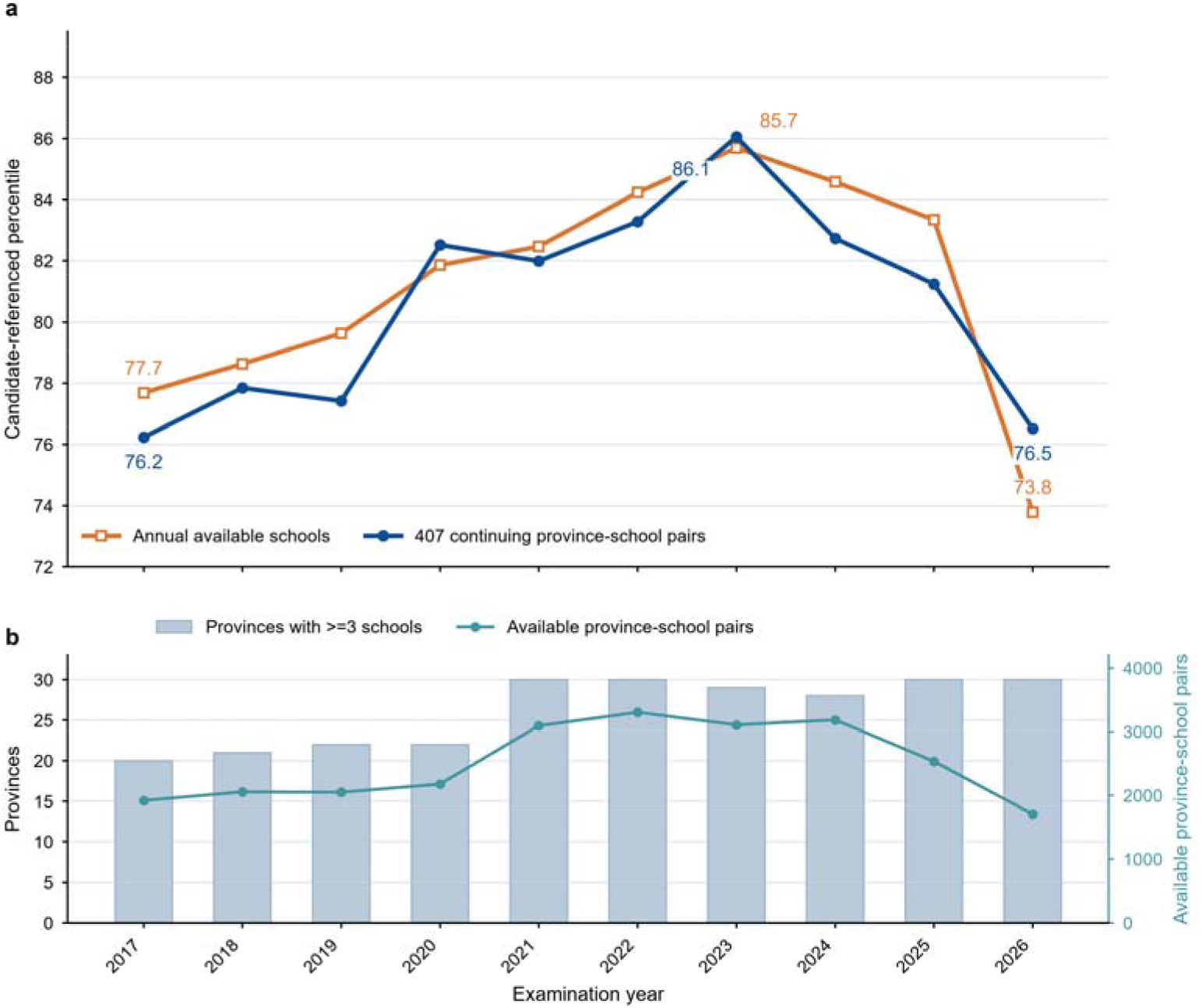
Annual clinical medicine cutoffs and source coverage. Panel a shows the equal-province mean of provincial medians for annually available schools and the 407 province–school pairs observed in all ten years. Higher percentiles indicate earlier examination ranks. Panel b shows annual counts of contributing provinces (bars, left scale) and province–school pairs (line, right scale).

In the annually available sample, mean provincial cutoff percentiles were 77.69 in 2017, 85.69 in 2023 and 73.79 in 2026. The corresponding fixed-panel means were 76.23, 86.06 and 76.51 (Figure 1). All 15 continuously observed provinces moved earlier during 2017–2023 and later during 2023–2026. Omitting one province at a time produced recent declines of 8.46–10.10 points. The fixed-panel maximum occurred in 2023 under every coverage threshold. The annually available sample peaked in 2020 only when provinces with a single school could contribute, a coverage-sensitivity specification. The pre-transition 2023–2025 comparison included 2,084 province–school pairs in 29 systems. Provincial medians moved later in 25 provinces and earlier in four (mean −2.31 points; 95% province-resampling interval −3.36 to −1.25). The post hoc 2023–2026 comparison included 1,345 province– school pairs in 29 systems. Its national average was the mean change in provincial median cutoffs: −5.65 points (95% province-resampling interval −7.76 to −3.50). Twenty-five provincial medians moved later and four moved earlier. When the same schools were required in all four years, 1,090 pairs remained across 26 provinces. Twenty-three provinces moved later and three moved earlier (mean −6.82).

The 2023–2026 direction was unchanged under alternative school-identity definitions. Original names produced 1,345 matched pairs in 29 provinces, with 25 provincial medians moving later and four earlier (mean −5.65 points). School codes alone produced 1,413 pairs and a mean change of −5.49 points. Code followed by normalised names produced 1,418 pairs and a mean change of −5.45 points. Code followed by source match names produced 1,423 pairs and a mean change of −5.43 points. Each definition preserved the same 25-to-four provincial split (Table S2). Restricting the analysis to standardised examination categories produced 1,344 pairs in 29 provinces, 25 later and four earlier, with a mean change of −5.66 points (Tables S3–S6). The maximum difference from the original-name estimate was 0.23 percentile points. Weighting each school median by its number of contributing programme records also preserved the reversal. The fixed-panel means were 81.48 in 2017, 89.06 in 2023 and 82.13 in 2026 (Table S10).

Figure 2 shows the provincial levels, missing years and cumulative changes.

**Figure 2.**
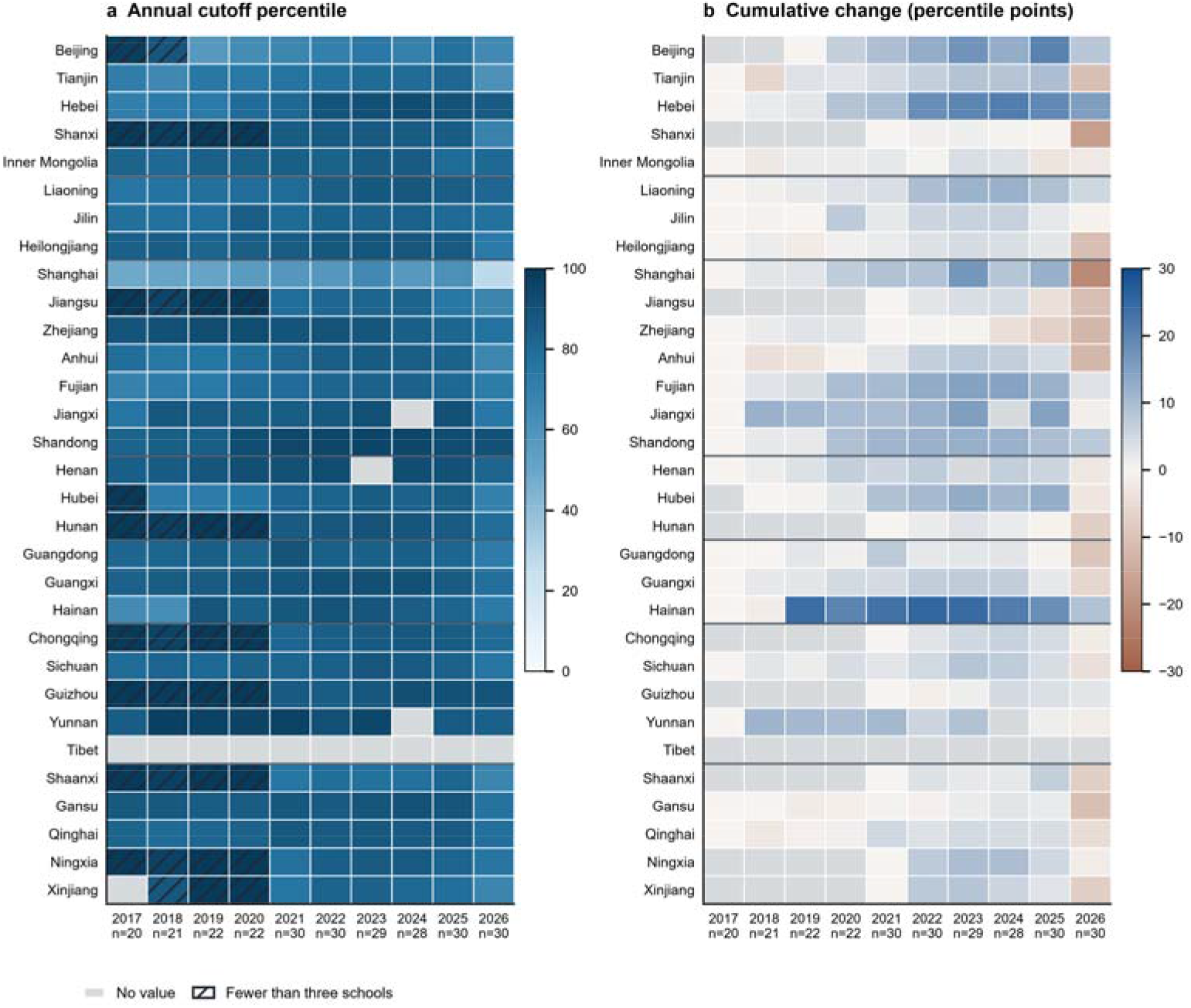
Provincial clinical medicine cutoff levels and cumulative changes. Provinces follow the standard administrative-region order: North, Northeast, East, Central, South, Southwest and Northwest. Horizontal rules mark the regional boundaries. Panel a shows the annual median cutoff percentile on a 0–100 scale. Panel b shows cumulative change in percentile points from the first year in which that province met the three-school threshold. Hatched cells indicate fewer than three schools; grey cells indicate no usable value. Higher percentiles indicate earlier examination positions, and negative changes indicate movement towards later positions.

### Measurement sensitivity

From 2017 to 2026, the province-equal matched-school change was +3.02 percentile points among 754 province–school pairs available at both endpoints. Including unpaired schools within those provinces next changed the estimate by −6.44 points. Expanding province coverage then changed it by −0.49 points, giving an annual-sample estimate of −3.90 (Figure 3, separately rounded components). Both samples moved later from 2023 to 2026: the matched estimate was −5.65 and the annual-sample estimate was −11.90. The 2023–2026 comparison is post hoc because 2023 was the observed maximum in the annual series. Full-decade estimates ranged from +2.71 to +3.02 across minimum school counts of 1, 3, 5 and 10. Each of the three recent-window estimates was stable across thresholds (Table S11). Sensitivity analyses that varied record deduplication retained the directions of the ten-year comparisons.

**Figure 3.**
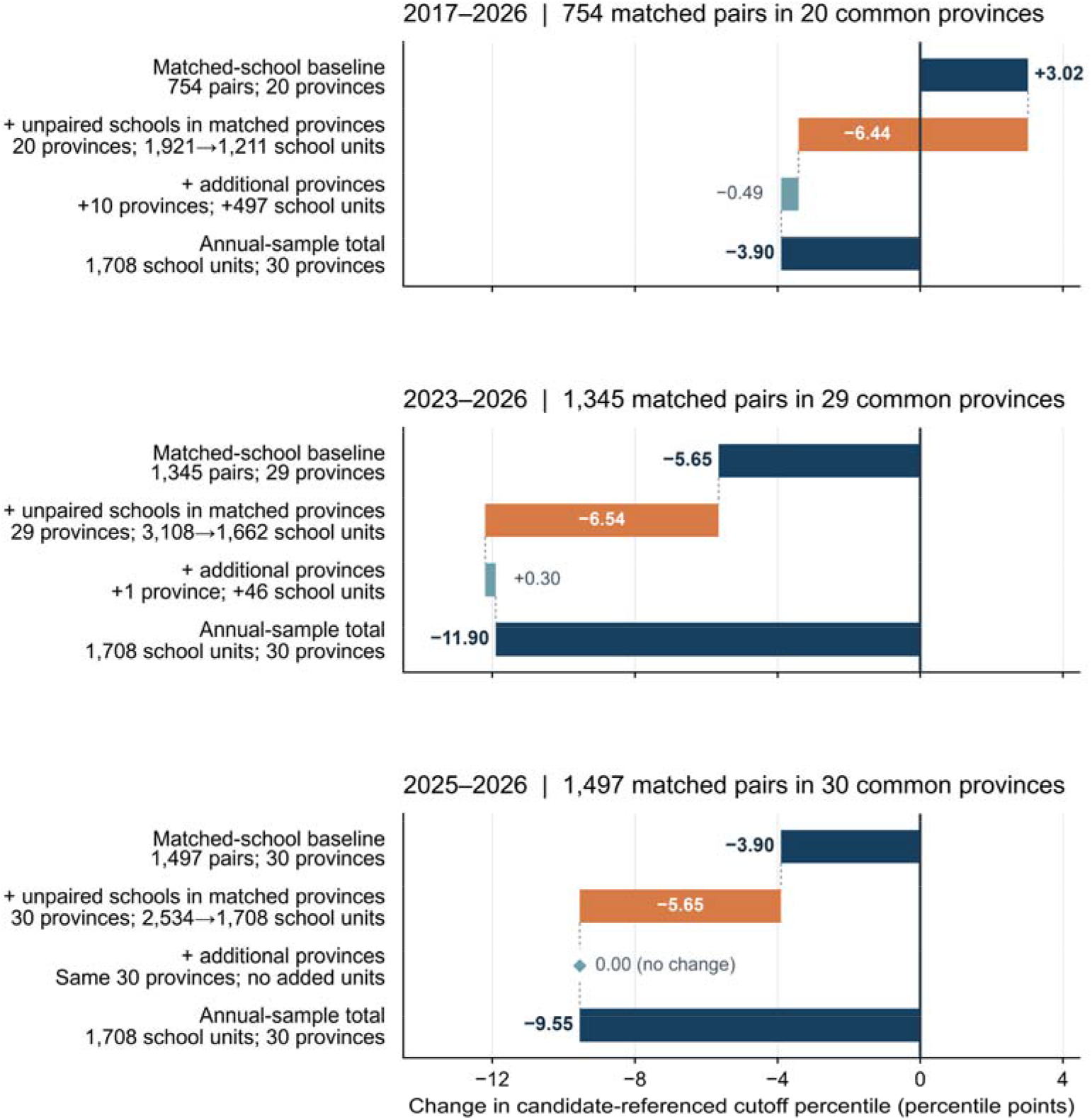
Sequential coverage-frame adjustments to estimated changes in candidate-referenced clinical medicine admission-cutoff percentiles. In each panel, the first and fourth bars show the province-equal estimates for schools matched at both endpoints and for the annually available sample. The middle bars show sequential changes from including unpaired schools within matched provinces and then expanding province coverage. These steps are sample adjustments that depend on their order. They should not be read as independent effects, causal contributions or corrections for nonrandom missingness. School-unit counts show the numbers of schools observed at the starting and ending endpoints; matched pairs are counted once at baseline. The 0.00 province-coverage adjustment in 2025–2026 indicates that no additional provinces entered at the final step and was zero by construction. Negative values indicate movement towards later candidate ranks. Components are rounded separately. Uncertainty intervals are not shown.

### Source and category sensitivity

Across nine adjacent-year comparisons, the two measures agreed in direction for 71.2%–85.8% of matched schools and 40.0%–81.5% of provincial medians (Supplementary Figure S1). In 2025–2026, 1,057 of 1,497 province–school pairs moved later on both measures. Another 228 moved earlier on both, 210 disagreed and two tied on the candidate measure. Across all 1,497 fixed pairs, 1,234 moved later, 261 earlier and two tied (school-pair mean −3.31 percentile points). For the same window, the province-equal mean of provincial median changes was −3.90 points (Figure 3). The dominant source family changed in 1,329 pairs. Of these, 1,096 moved later, 232 earlier and one tied (mean −3.38 points). The 168 pairs with an unchanged source family came from Shanxi, Zhejiang and Qinghai. Their school-weighted mean was −2.77 points and their province-equal mean was −2.32 points. All three provincial medians remained negative, providing geographically narrow corroborative evidence.

### Training pathways and reference populations

In 2023–2026, 25 provincial systems contributed at least three schools with a common training pathway. Only 126 of 1,046 matched province–school pairs had two or more shared pathways. Equal weighting of shared pathways yielded a 4.15-point decline, with 21 provincial estimates moving later and four earlier. The six-year subgroup is excluded from interpretation because the recent source-label audit found conflicting duration text. Across the ten cycles, the explicitly identified five-plus-three route usually reached an earlier rank than the five-year course at the same school and examination stratum. Median annual differences ranged from 0.43 to 1.74 points (694 comparisons in 29 systems).

In the examination-category analysis, growth in the reference population offset part of the rank-based decline (Table 1). The supplement gives field availability and source-level plan reconciliation for all 310 province-years and the 100-cell source audit.

**Table 1.** Admission cutoff changes under endpoint matching, shared training pathways and matched examination populations.

| Comparison and period | Contributing provinces and province–school pairs | Estimates in percentile points | Interpretation |
|---|---|---|---|
| Endpoint matching, 2023–2025 | 29 provinces, 2,084 pairs | −2.31 | Cutoffs moved later in 25 provinces and earlier in four before the main 2026 source transition. |
| Training pathway, 2017–2026 | 20 provinces, 739 pairs | +3.27 with all coded records, +3.27 for common routes, +3.27 with common routes weighted equally | Cutoffs reached earlier ranks in these endpoint-matched schools over the full decade. |
| Training pathway, 2023–2026 | 25 provinces, 1,046 pairs | −4.27, −4.11, −4.15 under the three main specifications | The six-year subgroup was excluded from interpretation because source records conflicted on programme duration. |
| Training pathway, 2025–2026 | 30 provinces, 1,488 pairs | −3.86, −3.86, −3.85 | Changes were similar across the three definitions in this window. |
| Examination population, 2023–2026 | 15 provinces, 784 pairs | −7.06 in total, comprising −10.02 from absolute ranks and +2.95 from reference-population size | Replacing the reference population offset part of the rank movement. |
| Examination population, 2025–2026 | 30 provinces, 1,492 pairs | −3.88 in total, comprising −4.27 from ranks and +0.39 from reference-population size | The matched examination-category measure moved later in 28 provinces. |

**Table 1 note.** Negative values indicate a later rank. Pathway comparisons include five-year courses coded from explicit or probable durations unless specified. Each row uses its own eligible sample. Components are rounded separately. The supplement gives the 31-system coverage matrix and results from the original programme filings.

### Endpoint source verification

For the 2026 endpoint, published provincial distributions reproduced the frozen candidate denominator for 1,085 of 1,771 programme records (61.3%) across nine examination systems. The reproduced records included 904 exact-score tie endpoints confirmed independently. They also included 171 Shandong records that reported rank without a score, two records assigned the top-threshold endpoint and eight records assigned an endpoint above the top threshold. The remaining 686 records retained frozen source metadata without an independent denominator check. The endpoint audit therefore counted programme records, whereas the national trajectory sample counted 1,708 province–school pairs after school-level aggregation. For the post hoc 2023–2026 comparison, requiring all 2026 records in a province–school unit to be independently verified retained 772 matched pairs in nine systems. All nine provincial medians moved later, with a province-equal mean change of −5.64 points (Table S12). At the 2023 endpoint, 2,150 of 2,165 records matched a unique exact score in a multi-year public compilation. This count included 15 Xinjiang records reproduced from a reprint. These checks assess existing reference denominators and add no observations to the national endpoint estimate.

One paired programme record illustrates the endpoint calculation. Xi’an Medical University published 524 places for its ordinary five-year clinical medicine programme in Shaanxi before preferences opened in 2023 and 517 in 2024. Both cohorts were filled. The minimum score rose from 508 to 526, while the tied cutoff moved from ranks 27,965–28,337 of 166,437 to 31,870–32,295 of 174,874 (top 16.80%–17.03% and 18.22%–18.47%). Programme applications were unavailable. This single programme checks the capacity–percentile relation and does not change the national estimate. ^24,25^

### National admission-plan source audit

A total of 100 province-year cells across 30 provincial systems lacked a verified initial provincial plan total. These cells were audited. The registry contained 441 candidate plan rows and 492 links to admission records. Thirty-seven candidate values matched the frozen admission count, four field conflicts were adjudicated and two pre-choice plan values had independent dates. One cell contained both a verified plan and admission value. No complete provincial plan total was constructed (Table S13).

An additional project-level audit covered 110 province-university-programme-year units at three institutions across 26 origin systems in 2023–2026. Every unit had a published plan and a minimum admission boundary. Actual admissions were explicit for 78 units and first-parallel-round admissions for 16. The remaining 16 had a published plan and minimum boundary but no usable admitted count. None provided a matched count of all first-programme choices at the same province-school-programme grain (Table S14). A separate school-wide count at Chengdu University in 2023 recorded 265 first-programme choices and 91 admissions without a province-specific denominator. It was not added to the panel. These records broaden the source audit for plans and boundaries. They do not establish a national capacity total or a national applicant-demand series.

## Discussion

### A late-decade reversal

At the 2017 and 2026 endpoints, the matched-school estimate for 754 province– school pairs was +3.02 percentile points, whereas the annual-sample estimate was −3.90. The direction of the full-period change therefore depended on school and province coverage. In the continuously observed 15-province panel, the observed maximum occurred in 2023, with a province-equal mean of 86.06, followed by 76.51 in 2026. In the post hoc 2023–2026 comparison, provincial medians moved later in 25 of 29 comparable provinces, with a mean change of −5.65 percentile points. This recent direction remained stable across school-identity definitions and within the 15-province matched examination-population subset.

In the 2023–2025 comparison, provincial medians moved later in 25 of 29 provinces, with a mean change of −2.31 points. This shows that the recent direction was already present before the dominant 2026 source transition, although other source, coverage and measurement influences cannot be excluded. The 2023–2026 magnitude was larger and remained coverage-sensitive. The 2026 fixed-panel mean was close to its 2017 level, concentrating the reversal in the later years of the series. The annual-availability estimate remained 3.90 points below its 2017 level. The near-stable 407-pair continuous panel and the +3.02 endpoint estimate for 754 pairs reflect different sample compositions, including 15 versus 20 provinces and different continuing-school requirements.

### Career conditions and the relative appeal of medicine

Applicants assess medicine before they experience medical work. Their expectations may draw on training length, physicians’ reported conditions and the credential requirements they anticipate when seeking a clinical post. Available records are dated, discontinuous and differently sampled. Together they identify conditions present during the decade in which reported cutoffs moved, without forming a continuous national series.

Long working weeks appeared in each separately sampled survey. A 2015 national tertiary-hospital survey recorded a mean of 50.5 patient-service hours per week. ^15^ A 2017–2018 national tertiary-hospital survey found that about one in five physicians considered leaving the profession. ^16^ A 2018 survey of 2,915 physicians in 48 public hospitals across six provinces found that 88.9% worked more than 40 hours per week and 35.3% worked more than 60 hours. ^26^ In a 2019 survey of 22,213 physicians at 144 tertiary public hospitals in all 31 provincial systems, mean working time was 9.62 hours per day and 66.54% worked more than eight hours per day. ^18^ A national online survey conducted in 2020 and reported in 2021 included 12,930 physicians across 28 provincial systems. It recorded 7.77 clinical hours per day, 1.47 research hours per day and 5.77 working days per week. ^19^ A 2024 survey of 1,324 physicians in four provincial-level regions reported a mean of 49.4 total weekly hours. ^20^ Repeated psychiatric-hospital surveys recorded 47.1, 53.0 and 50.3 hours per week in 2017, 2019 and 2021. ^27,28,29^ The estimands differed, but every sample reported a long-hours profile.

National hospital statistics show the same service intensity across the wider delivery system. In 2023, general hospitals recorded 7.2 outpatient visits and 2.2 inpatient bed-days per physician per working day, compared with 6.8 visits and 1.9 bed-days in 2022. The 2024 yearbook also supplied province-level hospital workload values for all 31 provincial systems. ^30^ These measures capture direct service volume and exclude teaching, research, administration and waiting time.

The only repeated relative-earnings series covered 2016–2019. Its source-defined earnings ratio, calculated from after-tax physician income and gross average urban employee earnings, was 1.54, 1.92, 1.50 and 1.48; the recorded ratio declined after 2017. ^17^ The two components use different tax bases. Three further records provide dated standalone benchmarks. Mean annual pay was RMB 108,334 in a 2017 payroll study of 178,622 licensed physicians across 21 provinces. ^21^ More than half of respondents reported monthly income below RMB 7,500 in the 2020 online survey. ^19^ Mean self-reported monthly income was RMB 8,245.1 in the 2024 four-region survey. ^20^ These records use different samples, periods and remuneration definitions and were not treated as a continuous income series.

The length and structure of training add another entry cost. Clinical medicine includes five-year, integrated five-plus-three, six-year and eight-year routes. In the 2021 and 2022 national entrant cohorts, 84.76% and 78.39% of respondents expected further study before employment. ^14,13^ Credential requirements varied across two distinct recruitment frames (Table S15, Supplementary Figure S2). ^8,9,10,11,12^ Position lines and advertised places were different units from separate recruitment frames, and no hospital-tier effect was estimated.

These records describe long and heterogeneous training pathways, high service loads, varied recruitment requirements and a source-defined relative-earnings ratio that declined after 2017. Together, these conditions support a hypothesis of lower perceived net returns from medicine. The late-decade admission shift could reflect how entrants and families weighed these conditions alongside security, status and service mission, but applicant perceptions were not measured. Admission plans, applicant pools, programme composition, eligibility rules and allocation rules could also have contributed.

A 2026 discrete-choice experiment surveyed 555 clinical master’s and doctoral graduates. Pre-tax monthly income (33.7% relative importance) and job location (24.0%) were the most important attributes, and workload, career development, permanent-establishment status and working environment also influenced choices. ^31^ A national registration cohort recorded at least one change of practice institution for 725,424 of 3,748,224 full-time licensed physicians between 2011 and 2021; registry exits included retirement and did not identify voluntary career change. ^22^ These findings map the career stage after admission and show which working conditions graduates value. They do not connect those preferences to the applicants whose reported cutoffs moved.

### Career intentions and public accounts

A 2020, 24-province survey conducted during the COVID-19 period recorded how 21,085 senior-high students described medicine. Among 11,121 respondents who did not intend to apply, lack of interest was selected most often, followed by violence risk (18.5%), workload (17.5%), long training (17.3%) and high responsibility (14.7%). Low income was selected by 3.6%. ^23^ In this non-applicant subgroup, 47.3% came from Sichuan. The most visible objections concerned the nature of the work and its risks, with long training and workload more prominent than low income.

These intentions were measured before application and allocation. The available application-stage evidence is limited to a 2017–2019 Beijing statistical series and individual programme archives; it shows that application pressure and admission boundaries can move together or diverge as places and allocation rules change. ^32^ The cutoff records a published filing-or-admission boundary, while surveys and application records describe earlier expectations and choices.

### Provincial divergence

The national direction contains substantial provincial variation. From 2023 to 2026, cutoffs moved later in nine of ten eastern, five of five central, eight of eleven western and three of three northeastern systems. The equal-province mean changes were −8.31, −6.59, −2.70 and −6.07 percentile points. The direction therefore appeared across all four broad regions, with different magnitudes.

Local school supply, the mix of examination categories, candidate migration, living costs and the composition of hospital posts can produce different regional combinations of opportunity and pressure. Provinces with a larger supply of medical places may record a later cutoff without an equivalent change in the number of applicants. Provinces with limited places can retain an earlier cutoff even as interest weakens. Migration between provincial systems adds a further source of variation. The present data cannot assign a regional mechanism or estimate how much each condition contributed.

### International admission systems and labour-market signals

Comparisons with other admission systems clarify what a cutoff can and cannot represent. Portugal and Denmark publish first-choice numbers alongside medical-programme admission boundaries, but the two stages do not move in lockstep. ^33,34^ Quotas, programme capacity and allocation rules intervene between expressed preference and the final boundary. A first-choice count records demand at an earlier stage; a cutoff records the published boundary at the filing or admission stage, rather than applicant demand itself.

Japan provides a long-run counterpoint. Medical-school selectivity increased from 1980 to 2017 during prolonged economic stagnation, and occupational stability was one proposed interpretation. ^35^ This account suggests a contextual hypothesis: when secure employment and predictable earnings carry a high premium, medicine can become more selective even under adverse economic conditions. A later weakening of that premium could accompany the opposite movement as alternative careers improve or confidence in the relative security of medicine declines. The Japanese case does not imply that the two countries share a common trajectory or that the proposed conditions caused the Chinese reversal.

Costs recorded in the United Kingdom and the United States provide parallel labour-market context rather than additional evidence on Chinese trends. In the United Kingdom, 42% of responding doctors in training reported heavy or very heavy workloads. In the United States, 70% of the 2025 graduating survey reported education debt, with a median of USD 215,000 among indebted respondents. ^36,37^ Chinese entrants face a long domestic training route and a recruitment market differentiated by institution. American entrants face a large financial entry cost. British trainees face sustained service pressure. In each setting, medicine competes with other careers on security, earnings, working conditions and the time required to establish a career.

International comparisons show that applicant preferences and final admission cutoffs are distinct quantities. They provide contextual hypotheses but do not identify the mechanisms underlying the changes observed in China. Admission institutions determine where any such change becomes visible: in first-choice counts, application ratios, interview selection or the final cutoff. The Chinese series uses published filing- or-admission boundaries across a large and institutionally diverse national system.

### Interpretation and research implications

The central contribution is a province-equal national account of how reported clinical medicine cutoffs changed over a decade and how that account depends on school and province coverage. The recent reversal survives within-school, school-identity and examination-category comparisons, while its magnitude remains coverage-sensitive. The full-period direction changes when coverage definitions change. Separately sampled records document long hours, long and heterogeneous training, a source-defined relative-earnings ratio that declined after 2017, varied recruitment requirements across two frames in 2025–2026, and hesitation among prospective students. These records provide context around the admission series but do not estimate the causes of the cutoff changes.

Provincial applicant counts, first choices and complete place totals would separate demand from capacity and score distribution. Repeated provincial measures of physician hours, earnings and recruitment would show whether conditions recorded in separate snapshots moved with the national admission series. Following applicants through medical school, postgraduate training and the first clinical post could test whether a later cutoff predicts later entry, exit or migration within the profession. A linked applicant-cohort design is the next step for testing whether and how these conditions enter that choice.

### Limitations

Thirty-one provincial systems entered the frame, 30 contributed to at least one annual estimate and 28–30 contributed in each year from 2021 to 2026. Tibet had no province-year cell with three schools, and Henan lacked a qualifying common-school comparison for 2023–2026. Programme composition, examination categories and source families changed within and between provinces, particularly in 2026. Subject-eligibility rules and admission-batch structures also changed across provinces during the study period and could alter the eligible applicant pool within a reported examination distribution. The annual sample contained 1,708 province–school pairs in 2026, compared with 2,534 in 2025. Fixed-school and category-level analyses preserved the direction of the recent result without treating the annual-sample difference as a demand estimate. Sources sometimes referred to filing and sometimes to completed admission, and stage was not recorded consistently. Part of the change may therefore reflect stage composition.

Independent source checks reproduced the reference denominator for 1,085 of 1,771 2026 programme records (61.3%) and matched 2,150 of 2,165 2023 records at a unique exact score. The reproduced count comprised 904 exact-score tie endpoints, 171 reported ranks from Shandong, two top-threshold endpoints and eight values above the published top threshold. The remaining records retained frozen source metadata and were not treated as directly reproduced.

The contextual evidence consists of single-year snapshots and short repeated cross-sections. Recruitment evidence comes from readable public schedules in leading hospitals or one national hospital group; advertised requirements describe vacancies and leave completed hiring unobserved. The physician surveys concentrate on tertiary public hospitals, and the psychiatric series is specialty-specific with changing hospital coverage. The 2020 survey covered 24 provincial systems, and non-applicant responses were concentrated in Sichuan. The 2017 payroll study and the repeated earnings surveys use different samples and remuneration definitions. International sources use different admission stages, institutions and sampling frames. These materials support interpretation of the national admission series and provide a comparative framework for future measurement. The domestic evidence-coverage table records the available denominator and principal limit for each contextual source family (Table S16).

## Conclusions

Clinical medicine admission cutoffs in China reached an observed maximum in the 15-province fixed panel in 2023 and subsequently moved later in most comparable provinces. This post hoc recent direction appeared under alternative school identities and within the matched examination-population subset, although its magnitude remained coverage-sensitive. The 2023–2025 comparison showed that the direction predated the main 2026 source transition. Across the full decade, the direction depended on school and province coverage. Long hours, a source-defined relative-earnings ratio that declined after 2017, long and heterogeneous training pathways, varied recruitment requirements across two distinct frames, and hesitation among prospective students describe the career context but do not identify the causes of the cutoff changes. International comparisons distinguish applicant preferences from final admission cutoffs and provide contextual hypotheses rather than causal evidence. Linking applicants, available places and career outcomes within the same cohorts is the next step for testing whether and how these conditions enter that choice.

## Ethics approval and consent to participate

This study used publicly available aggregate records and did not involve human participants or individually identifiable data. Institutional ethics review was not required.

## Consent for publication

Not applicable.

## Availability of data and materials

The data and analysis code that support the findings of this study are openly available in Zenodo at https://doi.org/10.5281/zenodo.24477787. The archive contains the national province-year panel, source manifest, analysis code and figure source data. The supplement contains the full province-year coverage and field matrices, endpoint source-locator archive, contextual evidence source cards, recruitment archive, search-coverage ledger, programme-source audit and the domestic evidence-coverage table.

## Authors’ contributions

Hang Zhou: Conceptualization, Data curation, Formal analysis, Investigation, Methodology, Visualization, Writing – original draft.

Xiuyuan Li: Conceptualization, Methodology, Validation, Supervision, Project administration, Writing – review and editing.

Both authors read and approved the final manuscript.

## Data Availability

The data and analysis code that support the findings of this study are openly available in Zenodo at https://doi.org/10.5281/zenodo.24477787.

## Acknowledgements

Not applicable.

