## Supplementary material for "Clinical medicine admission cutoffs in China, 2017-2026: a coverage-sensitive national longitudinal analysis": Figure_S1.pdf

### a Agreement by adjacent admission years

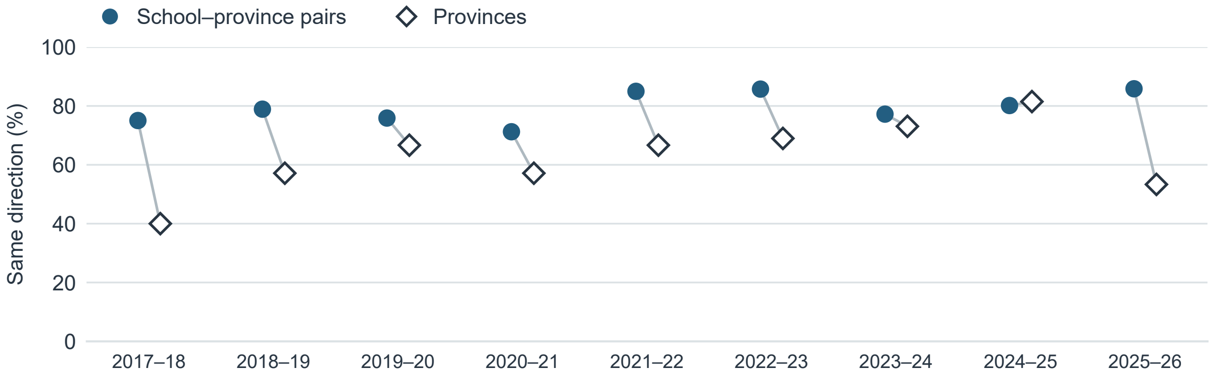


| | 2017–18 | 2018–19 | 2019–20 | 2020–21 | 2021–22 | 2022–23 | 2023–24 | 2024–25 | 2025–26 |
|---|---|---|---|---|---|---|---|---|---|
| School–province pairs | 1,691 | 1,844 | 1,866 | 1,964 | 2,768 | 2,732 | 2,537 | 2,085 | 1,497 |
| Provinces | 20 | 21 | 21 | 21 | 30 | 29 | 26 | 27 | 30 |

### b 2025–26 school–province pairs (n = 1,497)

| Candidate reference | Recorded programme-pool reference: Later | Earlier | Unchanged |
|---|---|---|---|
| Later | **1,057** | 177 | 0 |
| Earlier | 33 | **228** | 0 |
| Unchanged | 0 | 2 | 0 |

Spearman ρ for signed changes = 0.78

### c 2025–26 provinces (n = 30)

| Candidate reference | Recorded programme-pool reference: Later | Earlier | Unchanged |
|---|---|---|---|
| Later | **16** | 12 | 0 |
| Earlier | 2 | **0** | 0 |
| Unchanged | 0 | 0 | 0 |

Spearman ρ for signed changes = 0.19
