## Supplementary material for "Clinical medicine admission cutoffs in China, 2017-2026: a coverage-sensitive national longitudinal analysis": Figure_S2.pdf

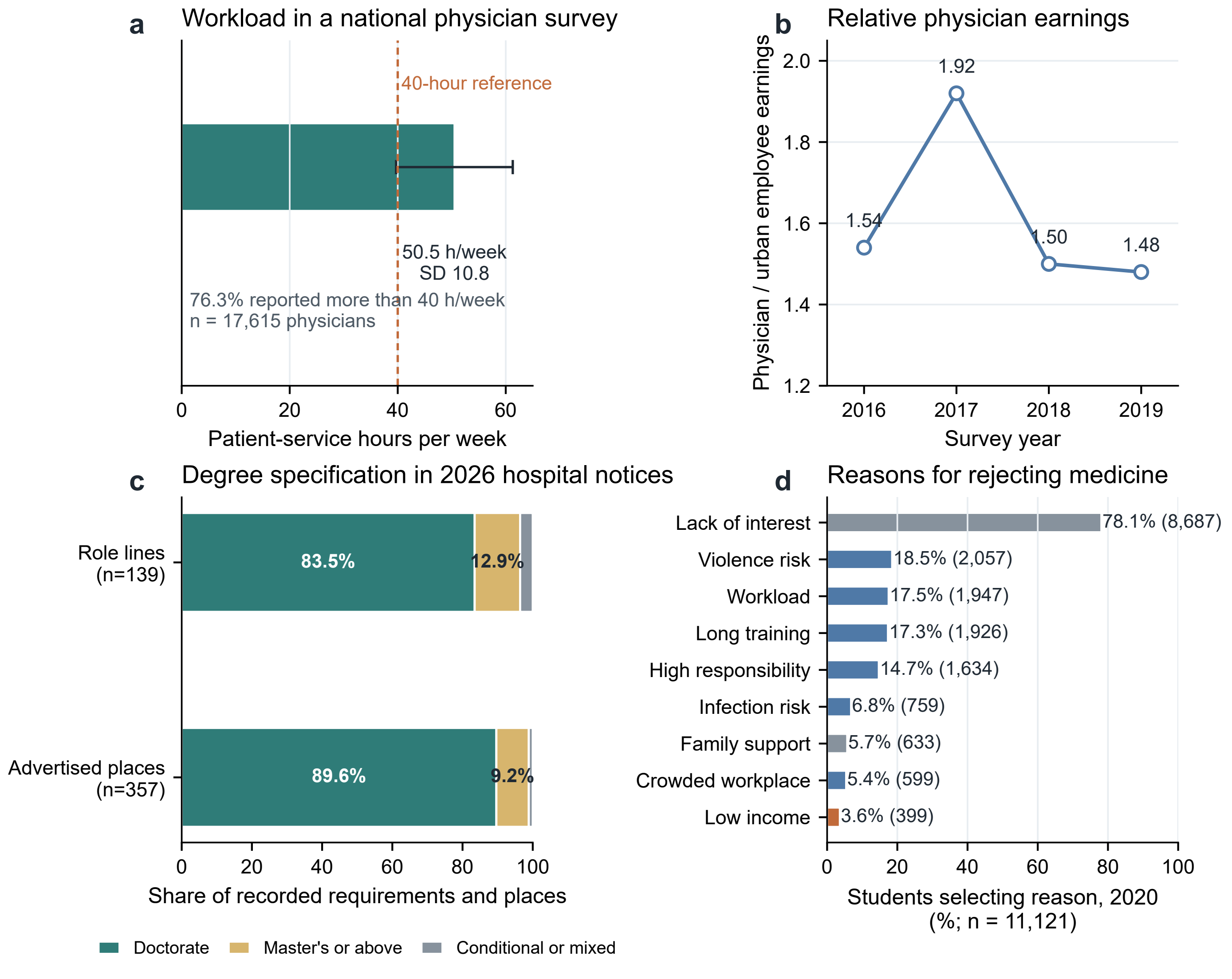
a
Workload in a national physician survey
40-hour reference
50.5 h/week
SD 10.8
76.3% reported more than 40 h/week
n = 17,615 physicians
0
20
40
60
Patient-service hours per week
b
Relative physician earnings
Physician / urban employee earnings
2.0
1.8
1.6
1.4
1.2
1.92
1.54
1.50
1.48
2016
2017
2018
2019
Survey year
c
Degree specification in 2026 hospital notices
Role lines
(n=139)
Advertised places
(n=357)
83.5%
12.9%
89.6%
9.2%
0
20
40
60
80
100
Share of recorded requirements and places
Doctorate
Master's or above
Conditional or mixed
d
Reasons for rejecting medicine
Lack of interest
78.1% (8,687)
Violence risk
18.5% (2,057)
Workload
17.5% (1,947)
Long training
17.3% (1,926)
High responsibility
14.7% (1,634)
Infection risk
6.8% (759)
Family support
5.7% (633)
Crowded workplace
5.4% (599)
Low income
3.6% (399)
0
20
40
60
80
100
Students selecting reason, 2020
(%; n = 11,121)
