## Supplementary figures and images for "Clinical medicine admission cutoffs in China, 2017-2026: a coverage-sensitive national longitudinal analysis"

### Figure_S1.png

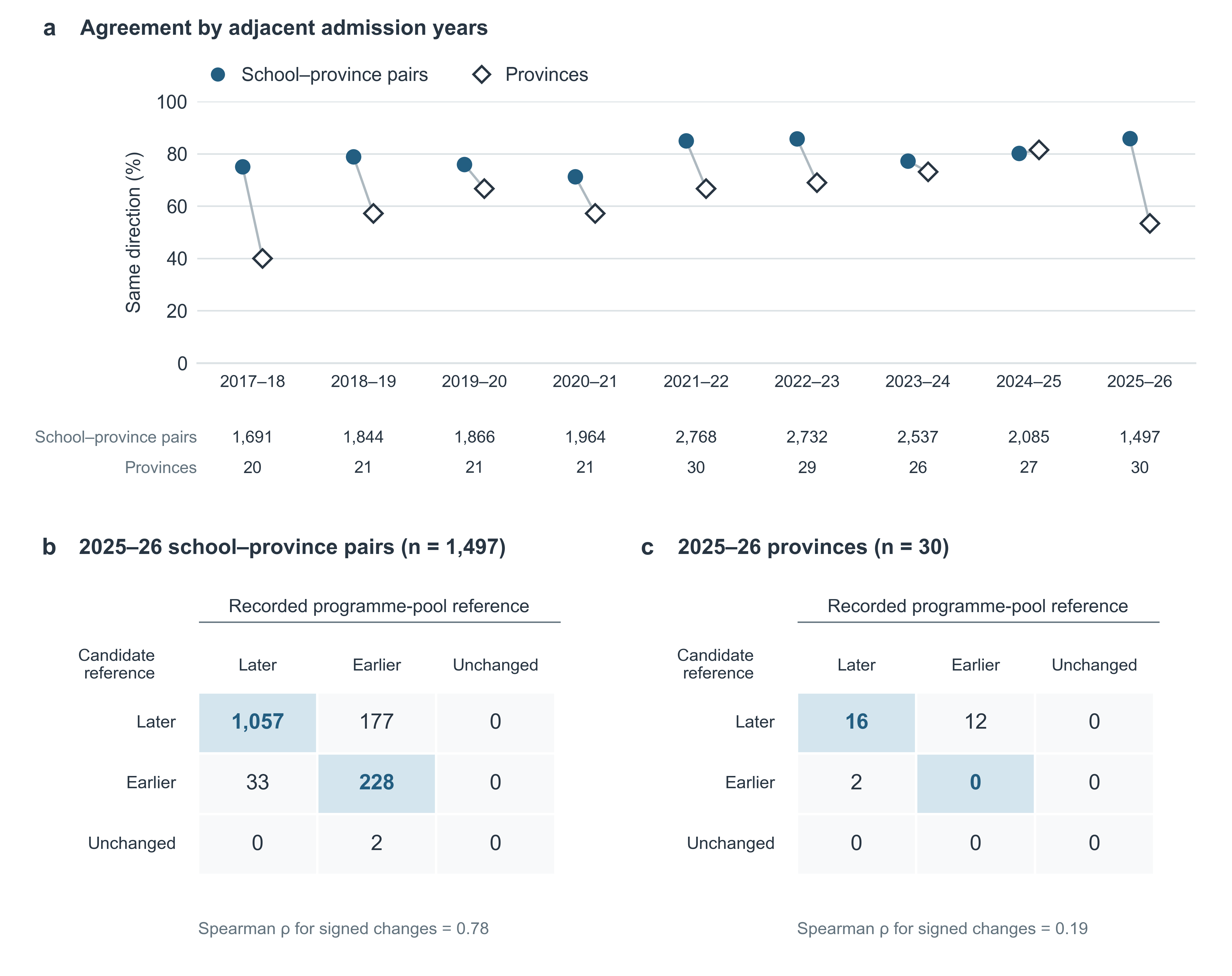

### Figure_S2.png

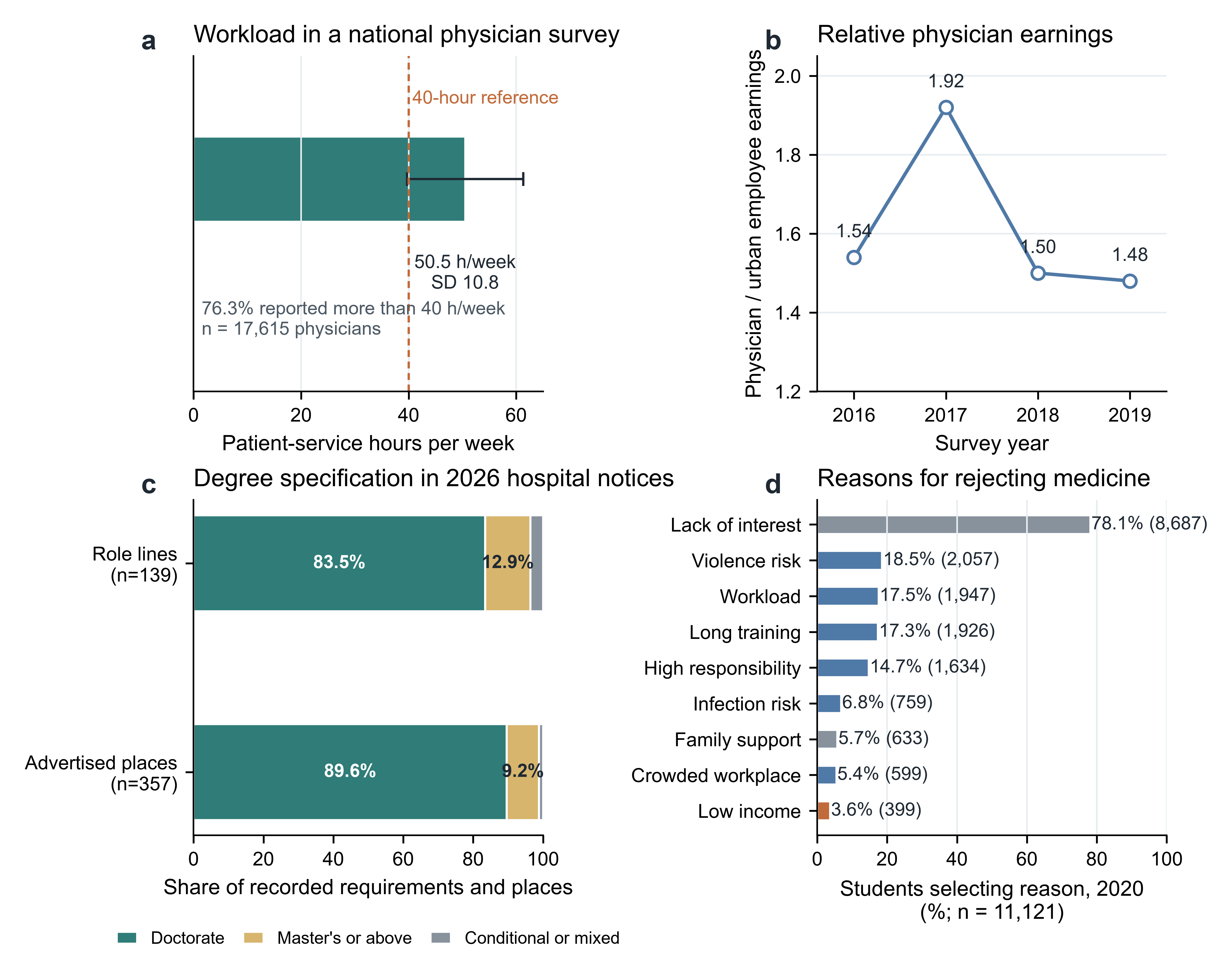
